# BREATH TEST TO DETECT WOMEN AT LOW RISK FOR BREAST CANCER: POTENTIAL CLINICAL AND ECONOMIC BENEFITS

**DOI:** 10.1101/2024.10.09.24315190

**Authors:** Michael Phillips, Therese B Bevers, Linda Hovanessian Larsen, Nadine Pappas, Sonali Pathak

## Abstract

**Background:** A breath test for volatile organic compounds has identified biomarkers associated with breast cancer. We evaluated the potential clinical and economic benefits of a breath test to detect women at low risk for breast cancer by comparing its negative predictive value (NPV) to the NPV of screening mammography.

**Methods:** Sensitivity and specificity values for screening mammography were obtained from the Food & Drug Administration Mammography Quality Standards Act; Amendments to Part 900 Regulations Docket No. FDA-2013-N-0134. The high values were sensitivity = 79.0%, specificity = 88.9% and the low values were sensitivity = 66.0%, specificity = 88.9%. In two previous studies of 771 women undergoing mammography, breath testing identified breast cancer with sensitivity=84% and specificity = 68.6% in 178 asymptomatic women, and sensitivity=82% and specificity = 77% in 593 who were symptomatic. These values were projected to a hypothetical screening population of 100,000 asymptomatic women with average breast cancer prevalence of 450/100,000, in order to estimate the NPV and PPV (positive predictive value) for breath testing and screening mammography respectively.

**Results:** Breath test in asymptomatic women: NPV = 99.895% and PPV = 1.19%; in symptomatic women: NPV = 99.895% and PPV = 1.59%. For screening mammography, NPV = 99.83% and PPV = 2.82% (low values), increasing to NPV= 99.89% and PPV = 3.12% (high values). A negative breath test identified 68.3% of the screening population as having low risk of breast cancer, with NPV similar to mammography. Based on Medicare reimbursement rates, elimination of mammography in women with a negative breath test could reduce the annual cost of breast cancer screening by 38.9%

**Conclusions:** In a hypothetical screening population, a negative breath test ruled out breast cancer with the same accuracy as a negative mammogram. A screening breath test could potentially eliminate the need for two thirds of all mammograms and reduce the costs of screening without increasing the risk of false-negative findings. If applied in clinical practice, this approach could potentially reduce the costs and burdens of breast cancer screening services, and benefit women by lessening the discomfort, anxiety, radiation exposure, and costs associated with mammography.

## INTRODUCTION

Mammography is one of the oldest diagnostic technologies in current use. The first reported mammogram was performed in 1913 by Dr. Albert Salomon, a surgeon in Berlin, who used an early X-ray machine to examine breast cancers^1 2^. Breast imaging technology has improved greatly during the past century, but its basic objective has not changed: to identify cancerous lesions in breast tissue. Screening mammography is widely regarded as the “standard of care” for detection of early-stage breast cancer, but it has achieved this status by default because of the lack of competing technology to challenge its role.

The main clinical limitation of mammography is its inefficiency as a tool for primary screening. In order to detect one case of breast cancer, radiologists need to perform screening mammograms in approximately 220 women, based on a prevalence of 450 cases per 100,000^***3-6***^. However, for the great majority of screened women (219/220 i.e. 99.6%), mammography functions solely as a “rule out” test for breast cancer. While this reassurance is important, it comes at a high cost: the discomfort of breast compression, anxiety, exposure to radiation, and financial burden. Women and their healthcare providers could benefit from a test to rule out breast cancer with the same accuracy as a mammogram, but without the associated discomfort, anxiety, radiation, and cost. This need has increased because access to screening mammography is limited by a growing shortage of radiologists and imaging services in the USA and other countries^7,8^.

Breath testing offers a new solution to an old problem: Mammograms and breath testing operate on fundamentally different principles: instead of imaging lesions in breast tissue, a breath test identifies volatile organic compounds (VOCs) exhaled in breath that are metabolic products associated with breast cancer. Several studies have identified distinctive breath VOCs in breast cancer patients using gas chromatography mass spectrometry (GC MS) ^*9 10*^ as well as nanosensor arrays^*11*^ and sniffing dogs^*12*^. We have reported a rapid point-of-care breath test employing gas chromatography and surface acoustic wave detection (GC SAW) to detect breath VOCs^*13*^. Breath VOC biomarkers were sensitive and specific for breast cancer in symptomatic as well as in asymptomatic women^*14*^. The biomarker VOCs were mainly alkanes and alkane derivatives that were consistent with products of increased oxidative stress ^*15-18*^. We report here an evaluation of the GC SAW point-of-care breath test as a “rule out” test on the bottom rung of the diagnostic ladder *(Figure 1)*, and the potential clinical and economic benefits of employing this strategy.

**Figure 1:**
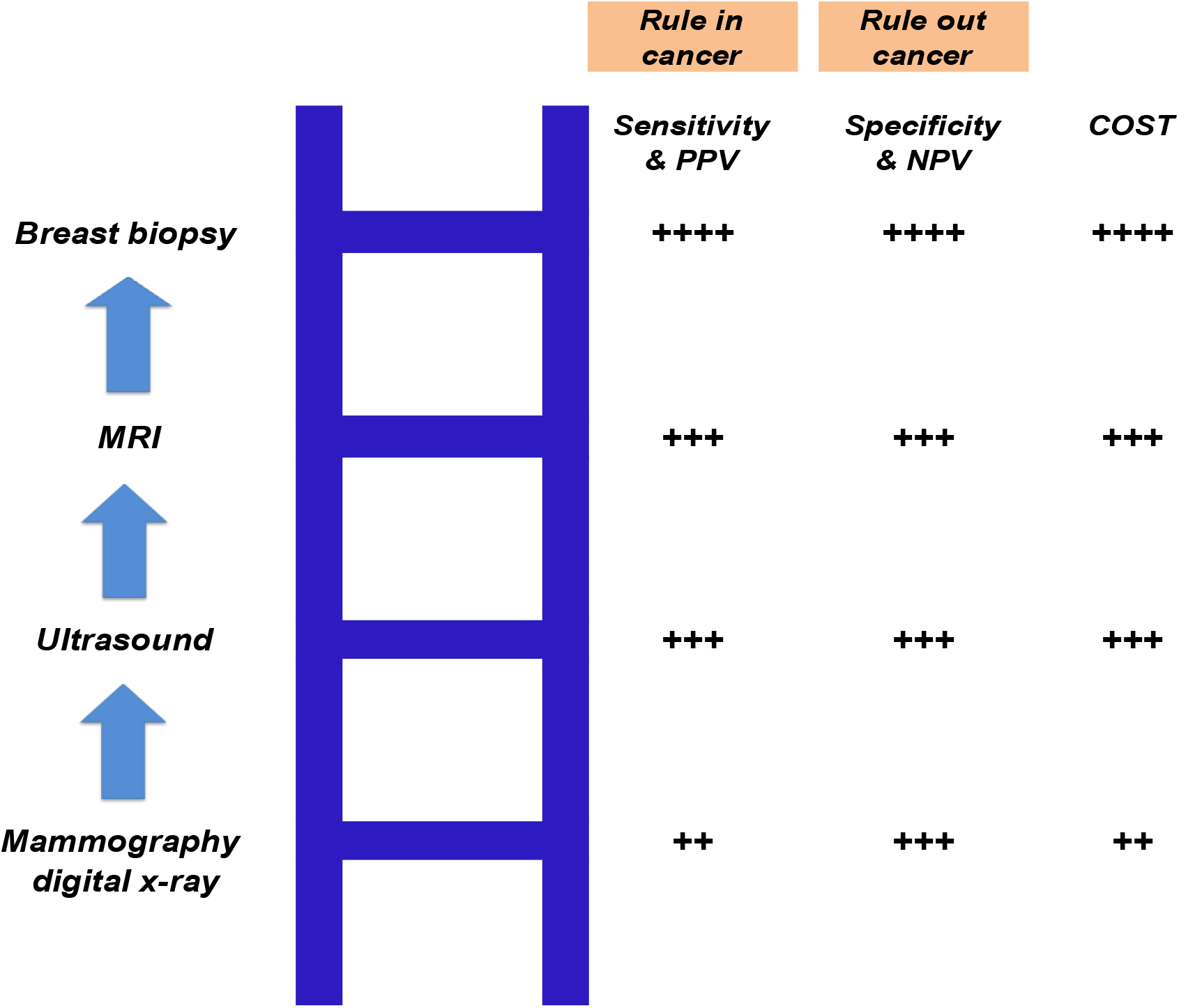
The diagnostic ladder of breast cancer screening. The diagnostic approach to breast cancer is analogous to climbing a ladder. The physician starts on the lowest rung of the ladder, usually with mammography employing a digital x-ray. If mammography is positive for breast cancer, the physician progresses to the next rung up the ladder. Since each test rung on the ladder has a high negative predictive value (NPV), a negative result will rule out breast cancer and no further testing may be needed. However, the positive predictive value (PPV) progressively increases with each test, culminating in the “gold standard” of breast biopsy on the top rung which has the highest sensitivity and specificity. The progressive increase in sensitivity and PPV on ascending rungs of the ladder is accompanied by increasing costs of testing, as well as increasing risks of potential harms to the patient.

## METHODS

### Sensitivity and specificity of tests for breast cancer

Values for screening mammography were obtained from the Food & Drug Administration (FDA) publication Mammography Quality Standards Act; Amendments to Part 900 Regulations Docket No. FDA-2013-N-0134^19^. According to FDA: “Results were shown from estimating annual values for screening mammography in the U.S.” and “Because data on sensitivity are difficult to obtain and estimates vary, calculations are presented using both a high and low estimate of sensitivity”. The high values were sensitivity = 79.0%, specificity = 88.9% and the low values were sensitivity = 66.0%, specificity = 88.9%. Breath testing with GC SAW was performed in two previous clinical studies of 771 women having screening mammography for breast cancer ^14^. Sensitivity=84% and specificity = 68.6% in 178 asymptomatic women, and sensitivity=82% and specificity = 77% in 593 who were symptomatic.

### Estimation of positive and negative predictive values (PPV and NPV) in a screening population

Studies of screening mammography performed in large populations indicate a representative prevalence of 450 breast cancers per 100,000 in asymptomatic women ^*3-6*^. This prevalence value was combined with the sensitivity and specificity values shown above in order to estimate the expected PPV and NPV of breath testing and screening mammography.

#### Estimation of economic effects in a screening population

Using the values shown in Fig 2, we estimated the costs of screening 100,000 women for breast cancer employing primary screening with mammography or a breath test respectively. Medicare recommends screening mammograms once every 12 months for woman aged 40 yr and older. The cost of a screening mammogram in the USA varies between $200 to $300 for uninsured women. The Medicare physician fee schedule reimbursement CPT code 77067 is used for bilateral screening mammography, including the use of computer-aided detection technology, The average Medicare-approved reimbursement for a diagnostic mammogram is around $170, and this value was employed as a conservative estimate of the annual cost of the test. The estimated average cost of a BreathX point-of-care test for breath VOCs employing GC SAW chromatography is $50.

**Figure 2:**
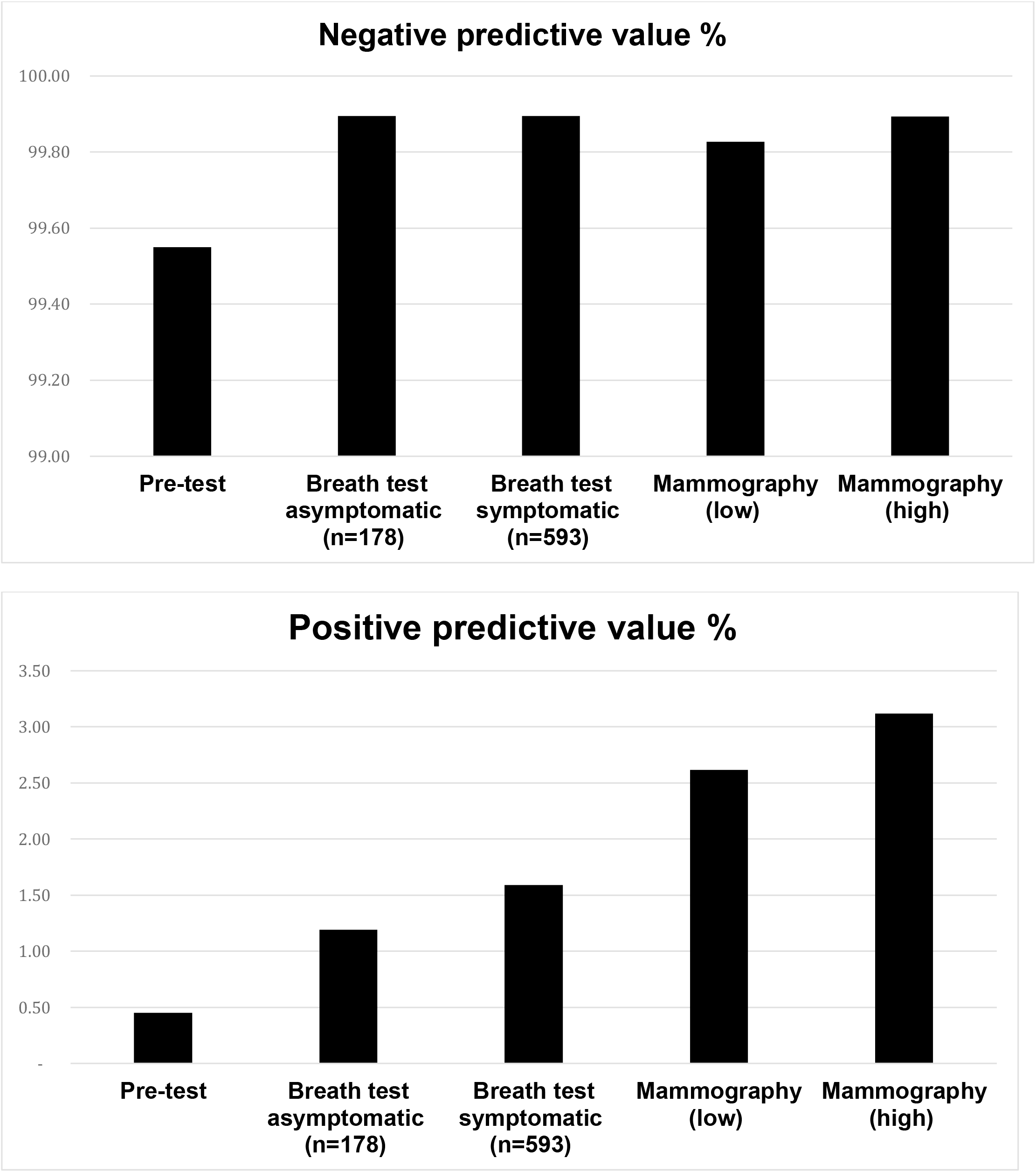
Positive and negative predictive values of breath test and mammography. This figure displays values of PPV (lower panel) and NPV (upper panel) derived from Table 1. Compared to pre-test values, breath testing and mammography both increased PPV and NPV. If the breath test is positive, a subsequent mammogram would increase the PPV and provide useful information about the likelihood of breast cancer. However, If the breath test result is negative, a subsequent mammogram would not increase the NPV, and it would not provide any additional diagnostic information to rule out breast cancer.

## RESULTS

### PPV and NPV of mammography and a breath test

Table 1 displays the expected outcomes of screening 100,000 asymptomatic women in a screening population. The outcomes of primary screening with mammography and a breath test respectively are shown in Figures 2 and 3.

**Table 1:** Positive and negative predictive values (PPV and NPV) of screening mammography and breath test. PPV and NPV were estimated in a representative sample of 100,000 asymptomatic women with an expected prevalence of 450 cases of breast cancer ^*3-6*^. Pre-test PPV and NPV represent the a priori values in the screening population. High and low values of screening mammography sensitivity and specificity were derived from FDA publication Mammography Quality Standards Act; Amendments to Part 900 Regulations Docket No. FDA-2013-N-0134^19^. Sensitivity and specificity of the breath test were derived from clinical studies of asymptomatic and symptomatic women employing a GC SAW breath test ^13 14^. Screening mammography had a higher PPV than a breath test, but NPV was similar with both tests.

|  |  | sensitivity% | specificity% | TRUE<br>POSITIVES | TRUE<br>NEGATIVES | FALSE<br>NEGATIVES | FALSE<br>POSITIVES | total | PPV% | NPV% |
| --- | --- | --- | --- | --- | --- | --- | --- | --- | --- | --- |
| **Pre-test** |  |  |  | 450 | 99,550 |  |  | 100,000 | 0.45 | 99.55 |
| **Post-test** |  |  |  |  |  |  |  |  |  |  |
| **Breath test** | asymptomatic (n=178) | 84 | 68.57 | 378 | 68,263 | 72 | 31,287 | 100,000 | 1.19 | 99.895 |
|  | symptomatic (n=593) | 82 | 77 | 369 | 76,753 | 81 | 22,797 | 100,000 | 1.59 | 99.895 |
| **Screening** | low | 66 | 89 | 297 | 88,500 | 153 | 11,050 | 100,000 | 2.62 | 99.827 |
| **mammography** | high | 79 | 89 | 356 | 88,500 | 95 | 11,050 | 100,000 | 3.12 | 99.893 |

**Figure 3:**
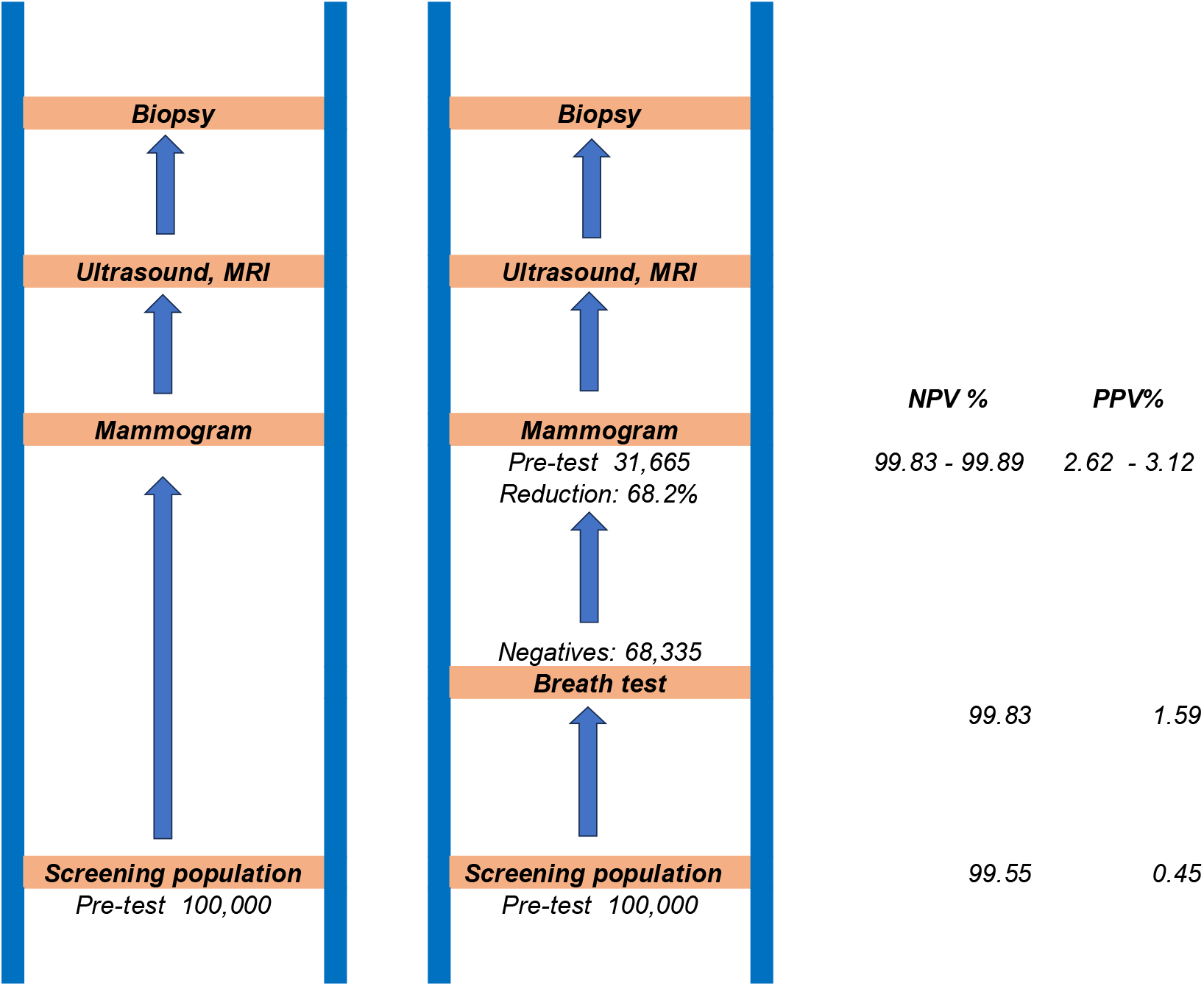
Effect of a breath test on the diagnostic ladder of breast cancer screening: This figure employs values for asymptomatic women derived from Table 1. The left-hand ladder displays the diagnostic ladder associated with primary screening of 100,000 women with mammography. The right-hand ladder demonstrates the expected outcome of employing a breath test as the first rung on the ladder. Since the NPV does not change, the number of women requiring a mammogram may be reduced by more than two thirds without incurring any increase in the false-negative rate

***Estimation of economic impact in a screening population*** is shown in Table 2. Employment of a breath test for primary screening followed by mammography for secondary screening reduced the overall cost of annual mammography by 38.9%.

**Table 2.**
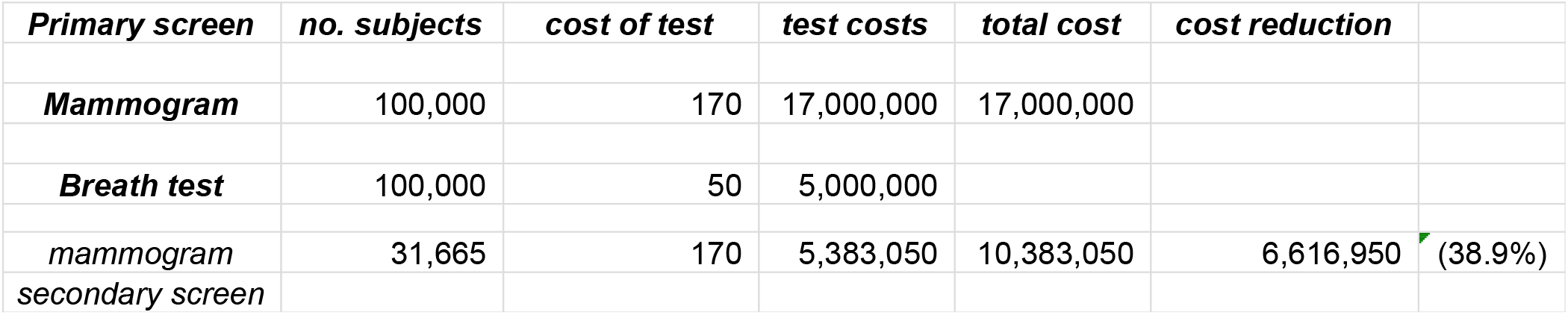
Estimation of economic impact of a screening breath test. This table shows the estimated costs of primary screening of a population of 100,000 with either mammography or a breath test. The numbers of subjects were derived from values shown in Table 1. The average Medicare-approved reimbursement for a diagnostic mammogram is around $170, based on Medicare physician fee schedule reimbursement CPT code 77067 used for bilateral screening mammography, including the use of computer-aided detection technology. For comparison, the average cost of a GC SAW breath test was estimated to be $50. All costs are shown in US dollars.

## DISCUSSION

The main finding of this study was that a breath test for VOCs identified women at low risk of breast cancer with the same accuracy as a screening mammogram. The NPV of a breath test (99.89%) was similar to the NPV of screening mammography (99.83 to 99.89%). Based on this finding, a breath test could potentially be employed in clinical practice for screening women as a “rule out” test for breast cancer with the same risk of a false-negative finding as a screening mammogram.

The clinical outcome of this approach is shown in Figures 2 and 3. The major potential benefit of primary screening with a breath test is that it could eliminate the need for more than two-thirds of all screening mammograms. As a result, the majority of women being screened for breast cancer would be spared most of the harms associated with mammography. They would suffer less discomfort and anxiety, and their exposure to radiation would be reduced.

The potential economic benefit of primary screening with a breath test would be a major reduction in costs to the healthcare system. Reduction in the number of mammograms could reduce the costs of screening for breast cancer by nearly 40%. As an additional benefit, screening with a breath test could potentially increase women’s access to breast cancer screening services by reducing the demands on overburdened radiologists and mammography centers.

Point-of-care breath testing is safe, painless and rapid, and it is as accurate as mammography as a diagnostic tool to identify women at low risk of breast cancer. A screening breath test for breast cancer biomarkers could provide major benefits for women by reducing their discomfort, anxiety, and radiation exposure. This approach could potentially benefit the healthcare system by reducing the costs of primary screening and improving access to overburdened screening centers.

## Data Availability

All data produced in the present work are contained in the manuscript

## Acknowledgements

Schmitt & Associates, Newark, NJ, and Michael Phillips analyzed the data. Breath test studies were funded by NIH NCI Grant Number: 5R44CA203019 – 02. ClinicalTrials.gov Identifier: NCT02888366.

## Notes

***Conflict of Interest:*** Michael Phillips is President and CEO of Menssana Research, Inc. All other authors declare that they have no conflict of interest.

### Competing Interest Statement

Michael Phillips is President and CEO of Menssana Research, Inc. All other authors declare that they have no conflict of interest.

### Clinical Trial

NCT02888366

### Funding Statement

The study was funded by NIH NCI Grant Number 5R44CA203019

### Author Declarations

Clinical studies were reviewed and approved by an Institutional Review Board (IRB) at all participating sites. Written informed consent was obtained from all individual participants. All procedures performed in studies involving human participants were in accordance with the ethical standards of the institutional and/or national research committee and with the 1964 Helsinki declaration and its later amendments or comparable ethical standards.

